# Towards a ML-powered Multiscale Computational Platform Based on QSP and PBPK Modeling to Support the Development of mRNA-based Therapies

**DOI:** 10.64898/2026.08.25.26361215

**Authors:** Elisa Pettinà, Frederick Abi Chahine, Elio Campanile, Stefano Giampiccolo, Luca Marchetti

## Abstract

mRNA-based therapeutics have emerged as a transformative class of medicines, yet their translation beyond infectious disease vaccines remains challenged by the absence of an integrated pharmacological framework accounting for the tri-component nature of these therapies - the lipid nanoparticle, the mRNA, and the expressed protein. Here, we present a modular, multiscale computational platform integrating two complementary mechanistic models covering the full pharmacological cascade of mRNA-based immunotherapies. The first is a Quantitative Systems Pharmacology (QSP) model describing the immunological response to mRNA vaccines, from antigen expression in antigen-presenting cells through B cell activation and circulating antibody production. The second is a Physiologically Based Pharmacokinetic (PBPK) model tracking whole-body disposition of mRNA-encoded therapeutic antibodies, incorporating a molecular layer resolving LNP uptake, endosomal mRNA escape, and intracellular translation. Both models are informed by a machine learning pipeline that maps IVT-mRNA nucleotide sequences directly onto kinetic parameters, enabling product-specific model simulations. We propose this platform as a step toward the quantitative pharmacological framework that mRNA therapeutics currently lack, and as a practical tool for model-informed design and development of this therapeutic class.

## 1 Introduction

Harnessing mRNA to instruct cells to produce a desired protein represents one of the most transformative advances in modern medicine. Decades of foundational research - spanning the discovery of nucleoside base modifications, the refinement of in vitro transcription (IVT) technology, and the development of lipid nanoparticle (LNP) delivery systems - converged in 2020 with the rapid development of mRNA-based COVID-19 vaccines [1,2].

To date, only two infectious diseases - COVID-19 and RSV - have approved mRNA-based vaccines, while mRNA therapeutics for other conditions, such as cancer and rare genetic disorders, remain in clinical development [3]. Advancing candidates to clinical trials remains hindered because of several reasons, and one of them is the lack of an appropriate pharmacological language. The existing pharmacological framework was designed for single-entity drugs, where the administered molecule is itself the therapeutic agent. mRNA-LNP therapies are fundamentally different: they are tri-component systems in which the lipid nanoparticle, the mRNA, and the expressed protein each exhibit distinct and interacting pharmacokinetic and pharmacodynamic behaviors [4]. Mathematical modeling offers a principled path forward: by explicitly representing each component and its interactions, it could provide the integrative, mechanistic framework that the field currently lacks [5].

With this goal in mind, we developed a platform of two complementary models supporting mRNA-based vaccines and monoclonal antibodies (mAbs), providing a unified modeling solution for *active* and *passive* immunotherapy, respectively. Our platform consists of a Quantitative Systems Pharmacology (QSP) model that represents the cascade of immunological events elicited by an mRNA-based vaccine, and a Physiologically Based Pharmacokinetics (PBPK) model that tracks the whole-body trafficking of mRNA-encoded antibodies (Figure 1). After fitting and validation on data retrieved from literature, we developed a Machine Learning (ML) pipeline that takes as input the IVT-mRNA sequence and directly informs mRNA-related parameters used in both our models.

**Figure 1:**
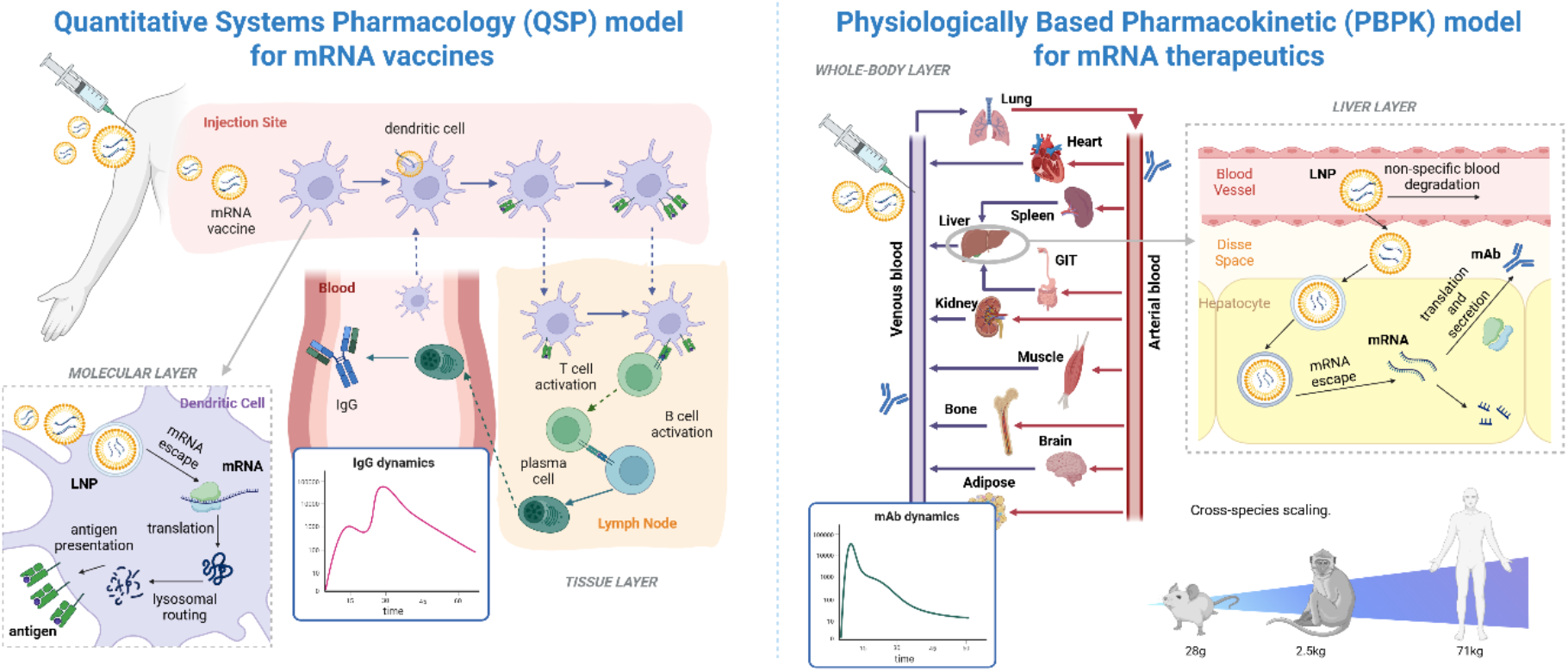
General overview of our computational platform. For the QSP model (left panel), both the tissue (describing the dynamics of antigen-presenting cells and the innate immunity activation) and the molecular layer (mRNA uptake and translation) are shown. Our PBPK model (right panel) comprises 14 compartments representing organs connected by the circulatory and lymphatic systems. The liver is where mRNA translation occurs, while the whole-body structure describes the PK of the therapeutics in the different organs.

Machine learning has shown considerable promise in drug development, and in the context of mRNA therapeutics, specifically for predicting sequence-dependent properties from the nucleotide sequence alone [6,7,8]. However, purely data-driven approaches produce statistical correlations that are difficult to interpret biologically and offer no insight into the mechanistic consequences of sequence-level differences. In our platform, a ML pipeline will bridge this gap by acting as a sequence-to-parameter translator: it converts mRNA design features into biologically interpretable quantities - translation efficiency and intracellular stability - that are directly propagated through the mechanistic layers of both models. This way, a less stable sequence is predicted to result in reduced mRNA availability, lower protein yield, and a consequently attenuated immune or pharmacokinetic response - making the biological implications of any sequence choice explicit and quantitatively traceable end-to-end.

## 2 Data and Methods

The QSP model, published in Dasti *et al*. [9], captures the key immune events triggered by mRNA vaccine administration, including tissue-level dynamics (antigen presentation, lymph node activation, effector cell recruitment) and a molecular layer describing LNP uptake into dendritic cells, endosomal mRNA escape, ribosomal translation, and antigen presentation.

The PBPK model extends our previous work [10,11] by adding a novel molecular layer at the hepatocyte level - the primary site of LNP-mediated mRNA delivery - tracking LNP uptake, endosomal escape, and translation into the therapeutic antibody, which then enters systemic circulation and is tracked via the two-pore/FcRn PBPK framework.

All data were obtained from published studies. The QSP model was calibrated stepwise: early antigen-presenting cell dynamics from NHP data [12], and adaptive immune parameters (T cell priming, B cell activation, antibody kinetics) from BNT162b2 clinical data [13,14,15,16,17], with mRNA-1273 fit using BNT162b2 as a starting point. Validation used independent dosing regimens and schedules for both vaccines [13,16,17,18,19,20,21].

The PBPK model was calibrated and validated on five public datasets of mRNA-encoded antibody pharmacokinetics (RiboMab02.1 [22], Pembrolizumab [23], B7H3×CD3 [24], Trastuzumab [25], XA-1 [26]), spanning different antibody formats, molecular weights (55-150 kDa), and LNP formulations, using recombinant-antibody data to estimate protein-specific parameters and mRNA-encoded data to estimate the molecular layer’s mechanistic parameters; validation used independent dosing and scheduling data from the same sources.

Despite performing well, both models lack the ability to predict the dynamics of new, unseen mRNA-based therapeutics. One reason is the mRNA molecule, whose half-life in the cytoplasm and translation rate can change dramatically. For this reason, we leverage two published neuronal-network (NN)-based tools, Saluki and RiboNN [6,8], and created a pipeline that programmatically takes in the mRNA sequence, converts it into FASTA format, and feeds it to the NNs. The final output will be an *ad hoc* estimate of the kinetic parameters 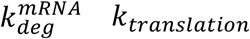,which, when inserted in our mathematical models, will allow a product-specific model simulation. In particular, the pipeline takes care of:

1. truncate the RiboNN neuronal network at the cell-type nodes to retrieve information specific to immune cells and hepatocytes;
2. calculate the predicted half-life of the mRNA from the score rank predicted by Saluki, attaching to the final node a regression model trained on known sequence-half-life data.

## 3 Results

The QSP model reproduced the immunological events following intramuscular administration of an mRNA-LNP vaccine, from antigen expression in APCs at the injection site through T cell priming, B cell activation in the draining lymph node, and circulating antibody production in blood (Figure 2). The model, calibrated on clinical RBD-binding IgG data from multiple independent cohort studies for the BNT162b2 (Pfizer-BioNTech) and mRNA-1273 (Moderna) vaccines, accurately reproduced circulating antibody time courses across different dosing levels and administration schedules [13,16,17,18,19,20,21]. Crucially, the model was not only able to fit observed data but also to identify, through in silico exploration, an optimal inter-dose scheduling regimen capable of maintaining continuous immune protection between administrations - a prediction that aligned with WHO recommendations. A key strength of the QSP component is its capacity to represent immunological heterogeneity. When the model was recalibrated using data from elderly cohorts [14,17], it quantitatively reproduced the attenuated antibody response characteristic of this population and provided mechanistic insight into the underlying biological differences - particularly in B cell maturation in germinal centers - that drive age-related decline in immune responses. Finally, the model’s cross-platform adaptability was demonstrated by applying it to the Moderna mRNA-1273 vaccine. Using parameter estimates derived from BNT162b2 calibration as a starting point, a single additional dataset at the 100 μg dose was sufficient to recalibrate the model, which was then successfully validated against independent data from the 25μg and 50μg dose levels, demonstrating that the platform’s mechanistic structure generalizes across mRNA vaccine products with minimal refitting effort.

**Figure 2:**
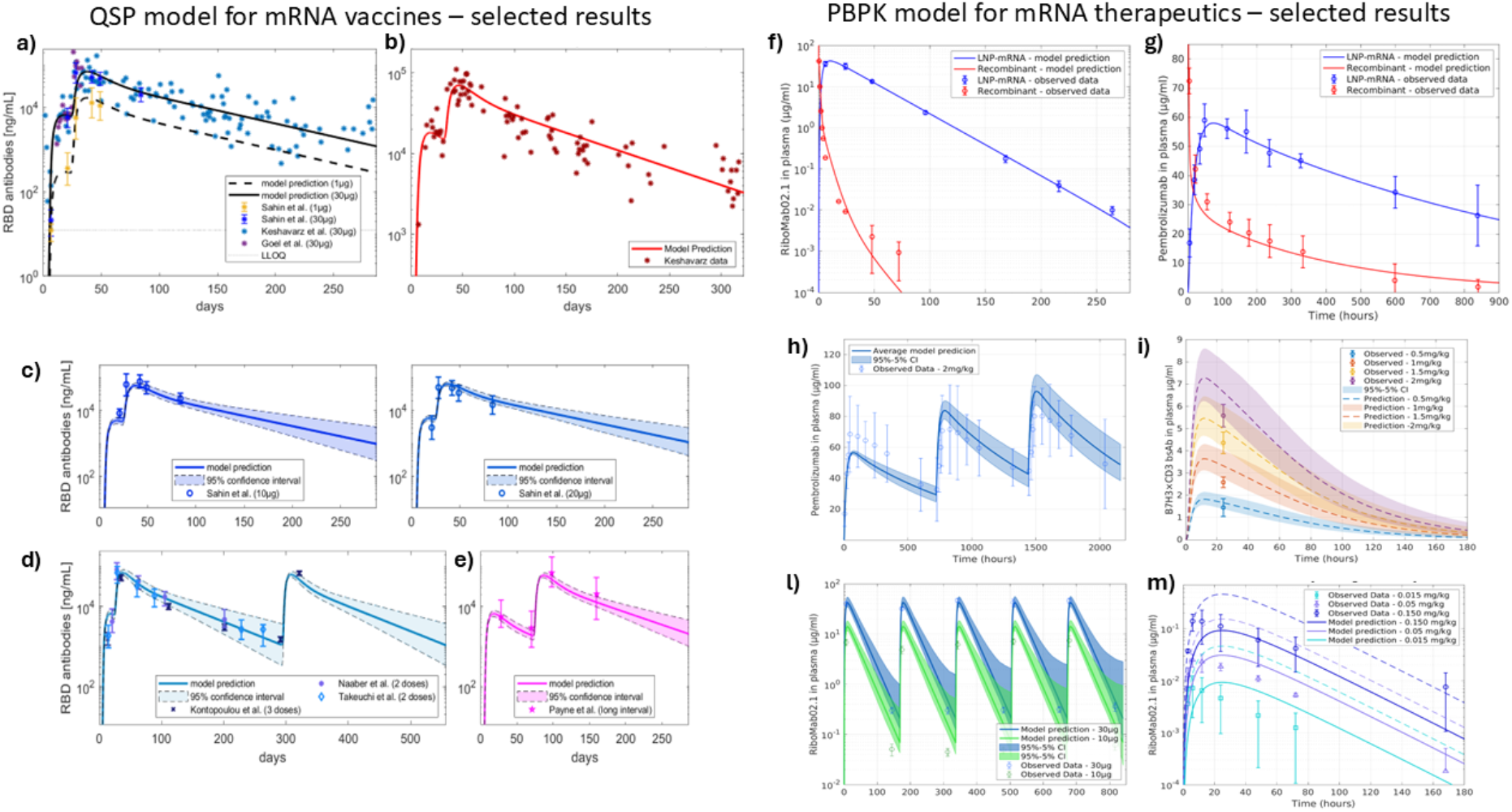
Selected results for the QSP and PBPK models. Left: QSP model calibration on a) PfizerBioNTech BNT-162b2 data, and b) Moderna mRNA-1273 data; QSP validation on c) 10μg and 20μg of the PfizerBioNTech BNT-162b2 data [13], d) three 30μg doses [16,17,18], and e) two 30μg doses administered with an extended dosing interval (10 weeks) [21]. Right: PBPK model calibration on f) RiboMab02.1 data [22], g) Pembrolizumab data [23]; PBPK validation on h) Pembrolizumab data [23], i) B7H3×CD3 bsAb data [24], l) RiboMab02.1 data (mouse) [22], and m) RiboMab02.1 data (NHP) [22].

The extended PBPK model, incorporating the hepatocyte-level molecular layer for LNP uptake, mRNA endosomal escape, and protein translation, successfully reproduced the pharmacokinetic time courses of mRNA-encoded therapeutic proteins and their recombinant counterpart (Figure 2). The molecular layer proved essential for accurately capturing the characteristic delay between LNP administration and peak therapeutic protein concentrations - a feature that arises from the sequential intracellular processes of endocytosis, endosomal escape, and ribosomal translation, and that cannot be reproduced by a standard one- or two-compartment pharmacokinetic model. Multi-dose validation in mice confirmed predictive accuracy across dosing schedules. The model was also successfully validated in NHP data for the RiboMab02.1 [22] product, with the caveat of increasing the LNP-mRNA degradation, a result that aligns with current literature findings [27].

The ML pipeline is being implemented and integrated into the platform architecture in both its modalities. By fixing the mRNA’s molecular kinetic parameter, the model results are more identifiable and reliable, and a step forward in modeling solutions that can be predictive, also for new therapeutic applications.

## 4 Conclusion

We presented a modular, multiscale platform integrating a QSP model of the mRNA vaccine-elicited immune response and a PBPK model of mRNA-encoded antibody disposition, providing a unified computational framework for both active and passive mRNA-based immunotherapy. Fitted and validated against clinical and preclinical data from BNT162b2, mRNA-1273, and a range of mRNA-encoded antibody formats, the platform enables mechanistically grounded predictions of dose-response relationships, inter-dose scheduling, and cross-species scaling. The architecture’s modularity allows individual components to be recalibrated for specific products and delivery routes. The integration of an ML pipeline for sequence-to-parameter mapping further extends the platform’s utility toward rational mRNA sequence design, with full validation currently underway. Together, these results position the platform as a practical tool for model-informed development of mRNA therapeutics and as a step toward the quantitative pharmacological framework that this therapeutic class currently lacks.

## Data Availability

All data produced in the present work are contained in the manuscript.

## Note

This paper has been accepted for the CIBB 2026 conference (https://cibb2026.teralab.ai).

